# Safety and protection of plasma donors: A scoping review and evidence (gap) map

**DOI:** 10.1101/2023.07.12.23292560

**Authors:** Natalie Schroyens, Tine D’aes, Emmy De Buck, Susan Mikkelsen, Pierre Tiberghien, Katja van den Hurk, Christian Erikstrup, Veerle Compernolle, Hans Van Remoortel, the SUPPLY consortium

## Abstract

**Background and objectives:** As part of a large-scale European project aiming to safely increase plasma collection in Europe, the current scoping review identifies the existing evidence (gaps) on adverse events (AEs) and other health effects in plasmapheresis donors, as well as factors that may be associated with such events/effects.

**Materials and methods:** We searched 6 databases and 3 registries. Study characteristics (publication type and language, study design, population, outcomes, associated factors, time of assessment, duration of follow-up, number and frequency of donations within the study period, convalescent plasma (y/n), study setting, and location) were charted in duplicate and in consultation with a content expert group. Results were synthesized narratively and in an interactive evidence gap map (EGM).

**Results:** Ninety-four research articles and 5 registrations focused on AEs (n = 38) and/or other health effects (n = 77) in plasmapheresis donors. Around 90% were observational studies (57 controlled; 33 uncontrolled), and most of them were performed in Europe (55%) or the USA (20%). Factors studied in association with donor health included donor characteristics (e.g., sex, age) (n = 27), cumulative number of donations (n = 21), donation frequency (n = 11), plasma collection device or program (n = 11), donor status (first-time versus repeat) (n = 10), donation volume per session (n = 8), time in donation program (n = 3), preventive measures (n = 2), or other (n = 9).

**Conclusion:** The current scoping review and EGM provide accessible tools for researchers and policy-makers to identify the available evidence and existing research gaps concerning plasmapheresis donation safety. Controlled, prospective studies with long-term donor follow-up are scarce. Furthermore, additional experimental studies comparing the health effects of different donation frequencies are required to inform a safe upper limit for donation frequency.

## Introduction

Plasma-derived medicinal products (PDMPs) are essential for the prophylaxis and treatment of several disorders. The vast majority of plasma used for the manufacturing of PDMPs stems from source plasma from plasmapheresis donations. During this procedure, plasma is retained whereas blood cells are returned to the donor ^1, 2^.

The production of PDMPs heavily relies on plasma that is collected outside of Europe, with the US providing the largest source of human plasma globally ^1, 3^. Such dependency entails a considerable risk of shortages, particularly during health crises such as COVID-19. A scarcity of source plasma, and thus of PDMPs, would threaten the supply of essential pharmacological treatments. In addition, safety regulations regarding plasma (donation), such as donor reimbursement and max. donation frequency, differ between Europe and USA. Thus, plasma collection in the EU should be expanded to ensure a stable and adequate supply of PDMPs. However, evidence-based guidelines on increasing plasma collection while maintaining plasma and donor safety are currently lacking.

With the ultimate aim of preparing recommendations for blood establishments, competent authorities, and medical societies for safely increasing plasma collection, the European Blood Alliance (EBA) initiated the ’SUPPLY’ project (Strengthening voluntary non-remunerated plasma collection capacity in Europe) ^4^. Situated within the SUPPLY project, the current scoping review aimed to systematically identify and map the available evidence (gaps) regarding adverse events (AEs) and other health effects in plasmapheresis donors ^5^. In addition, factors that have been studied in association with AEs or health effects are discussed, with a particular focus on donor status (first-time versus repeat donation), cumulative number of donations, donation frequency, and preventive measures.

## Methods

This scoping review is reported according to the ’PRISMA Extension for Scoping Reviews’ guideline ^6^ (Appendix 1).

### Preregistered study protocol

The preregistered study protocol (https://osf.io/hqj6z) and an overview of deviations (https://osf.io/8zx62) from the protocol were published on the Open Science Framework (OSF) ^7^.

### Eligibility criteria

#### Population

Include: adults who underwent plasma withdrawal via plasmapheresis.

Exclude: combination of donors giving either plasma or e.g., blood, platelets, … without separate data per donor type, donors giving multiple products at once (e.g., concurrent donation of platelets and plasma via plateletpheresis), recipients of plasma (-derivates), and patients.

#### Concept and context

Include: safety of plasma donation(s) in adults, regardless of the setting and geographical location of data collection.

Exclude: donor recruitment or retention, effects of plasma processing and fractionation, plasma safety for the recipient, supply of plasma(-derivates).

#### Outcomes

Include: [1] adverse events (AEs), defined and categorized according to the ’Standard for Surveillance of Complications Related to Blood Donation’ ^8^ with two additional categories: ’generic’ (i.e., study mentioning the measurement of ’AEs’ as such, without further specification or classification) and ’other’ for AEs not covered by the existing categories and [2] other health effects, including physiological parameters (e.g., blood pressure, heart rate), and parameters measured in blood or plasma (e.g., the concentration of blood cells, proteins, electrolytes).

Exclude: health-related outcomes that cannot clearly be linked to plasma donation (including self-reported general health status, measurements in the plasma product after donation without analysis of a possible link with donor/collection characteristics, or when the product of plasma donation underwent processing steps before analysis).

#### Study design

Include: systematic reviews, (non-)randomized controlled trials, (un-)controlled observational studies, and narrative reviews (not included in data charting table but used as a source of potentially relevant studies).

Exclude: animal, e*x vivo,* or *in vitro* studies.

#### Publication Type

Include: registrations, study protocols, and peer-reviewed articles regardless of publication status, language, and date.

Exclude: conference abstracts, book chapters, editorials, dissertations, and letters to the editor.

#### Search strategy and study selection process

We searched for eligible studies on 10 October 2022 using the 6 databases, 3 registries, and search strings listed in Appendix 2. References were screened in duplicate (NS and HVR) using

EndNote X9. Disagreements were resolved by discussion between the reviewers and, if necessary, by consulting the expert panel (KvdH, CE, SM, PT, VC). A PRISMA flowchart was created by means of a Shiny app ^9^.

#### Data charting, synthesis, and presentation

Study characteristics (publication type and language, study design, population, outcomes, associated factors, time of assessment, duration of follow-up, number and frequency of donations within the study period, convalescent plasma (y/n), study setting, and location) were charted in duplicate. The categories and variables for which data were sought, were continuously adapted during the review process, as we followed an exploratory approach ^10^. Extracting study results, judging the quality of evidence, and answering specific research questions were outside the scope of this review ^5^ and were performed in a separate systematic review project ^11^. Extracted information, available at https://osf.io/h8js5, was synthesized narratively using frequency counting per conceptual category and an evidence gap map (<u>EGM</u>) was created using EPPI-Mapper ^12^.

## Results

### Search results

Figure 1 illustrates the review process. Two full texts could not be retrieved ^13, 14^. To avoid duplication of studies, the protocol ^15^ and two of the registrations ^16, 17^ of studies that are described by identified research articles ^18, 19^, are not included in our summary. The remaining 94 research articles and 5 registrations are described below.

**FIGURE 1.**
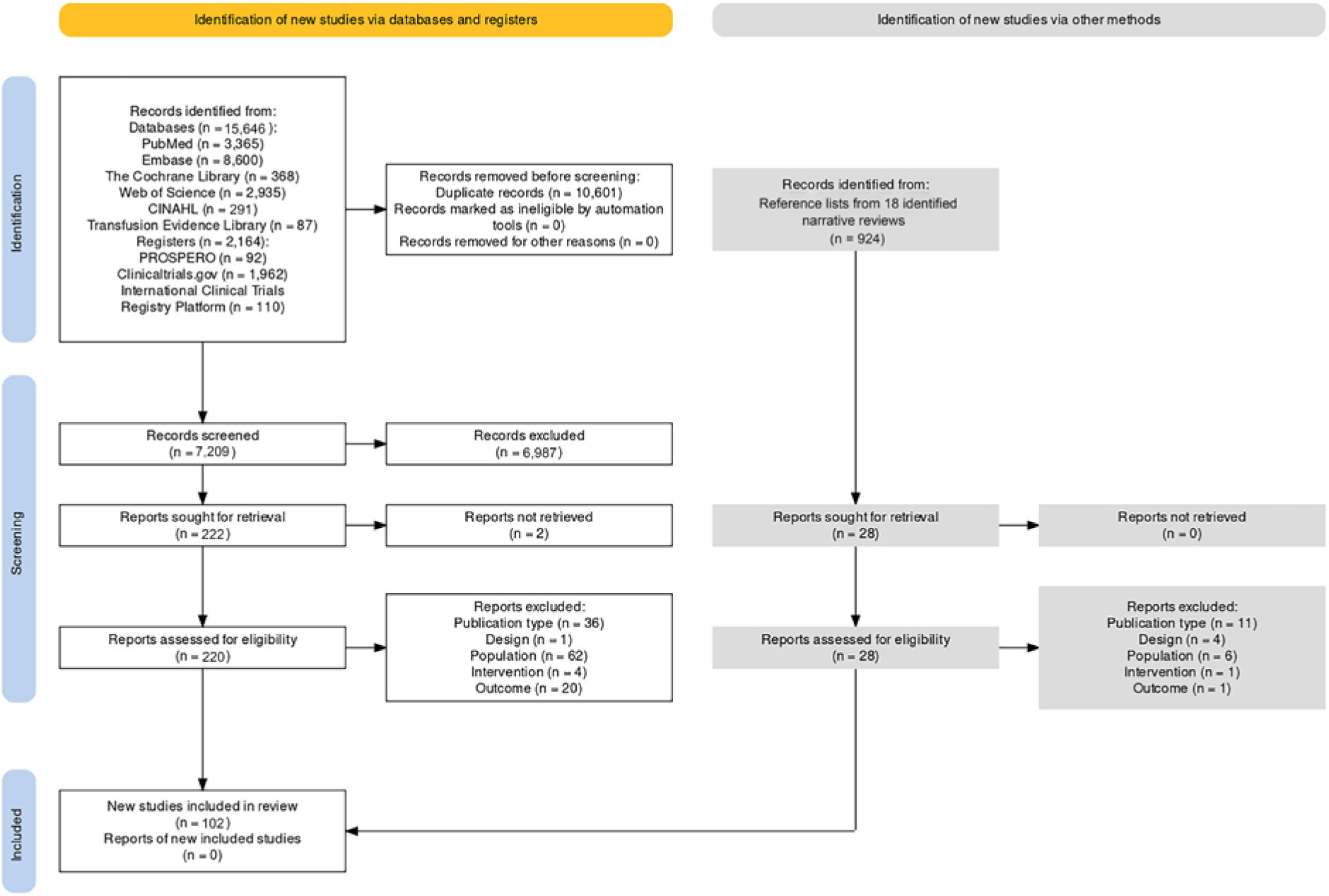
PRISMA Flow Diagram. Apart from 97 records identified via database and registry searches, 5 research articles were found via the reference lists of 18 narrative reviews (see https://osf.io/8zx62 for references). Overall, 102 records were obtained, including 94 research articles, 1 study protocol, and 7 registrations.

### Characteristics of included studies

#### Data charting table and evidence gap map

A detailed overview of study characteristics is available on OSF (https://osf.io/h8js5). Based on this overview, a more comprehensive and interactive evidence gap map (<u>EGM</u>) was created (see also Fig. 2-4) ^7^.

**FIGURE 2.**
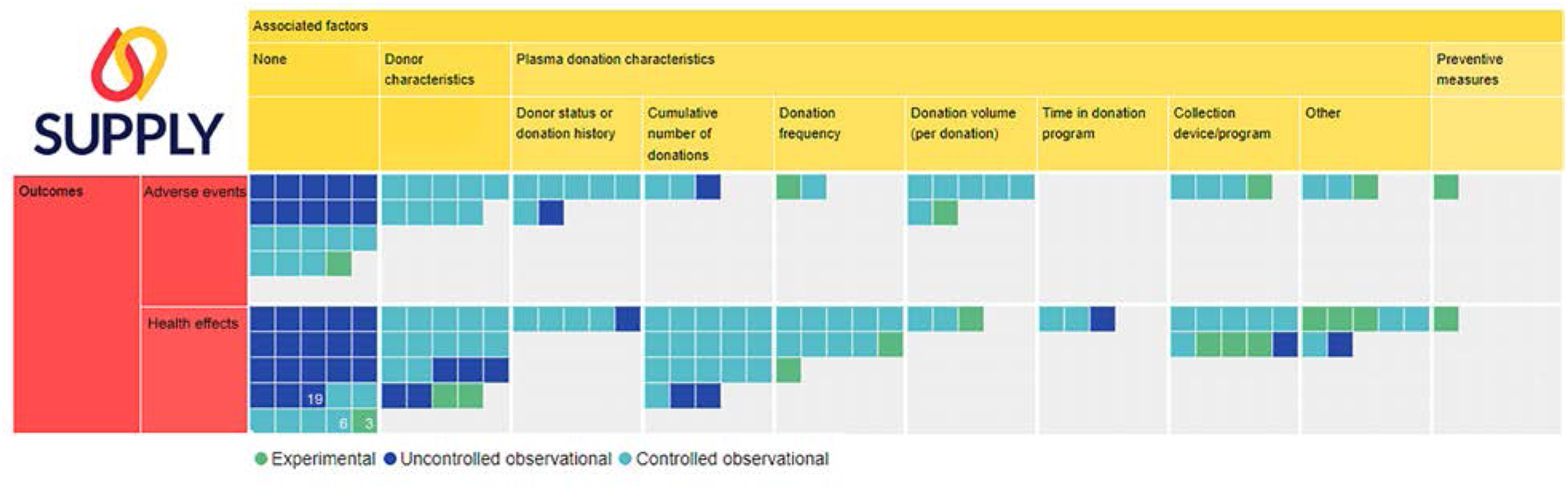
Evidence gap map, with rows representing outcomes and columns containing factors that have been studied in association with those outcomes. Squares represent studies, subdivided according to study design. An interactive version of the map is available <u>online</u>, in which descriptions of categories can be obtained by hovering over the column/row headers, lists of studies within each category can be obtained by clicking on the map, and studies can be filtered based on study design, publication type, publication date, location, publication language, follow-up period, population, and whether or not convalescent plasma was donated. Generated using v.2.2.4 of EPPI-Mapper powered by EPPI Reviewer and created by the Digital Solution Foundry team.

**FIGURE 3.**
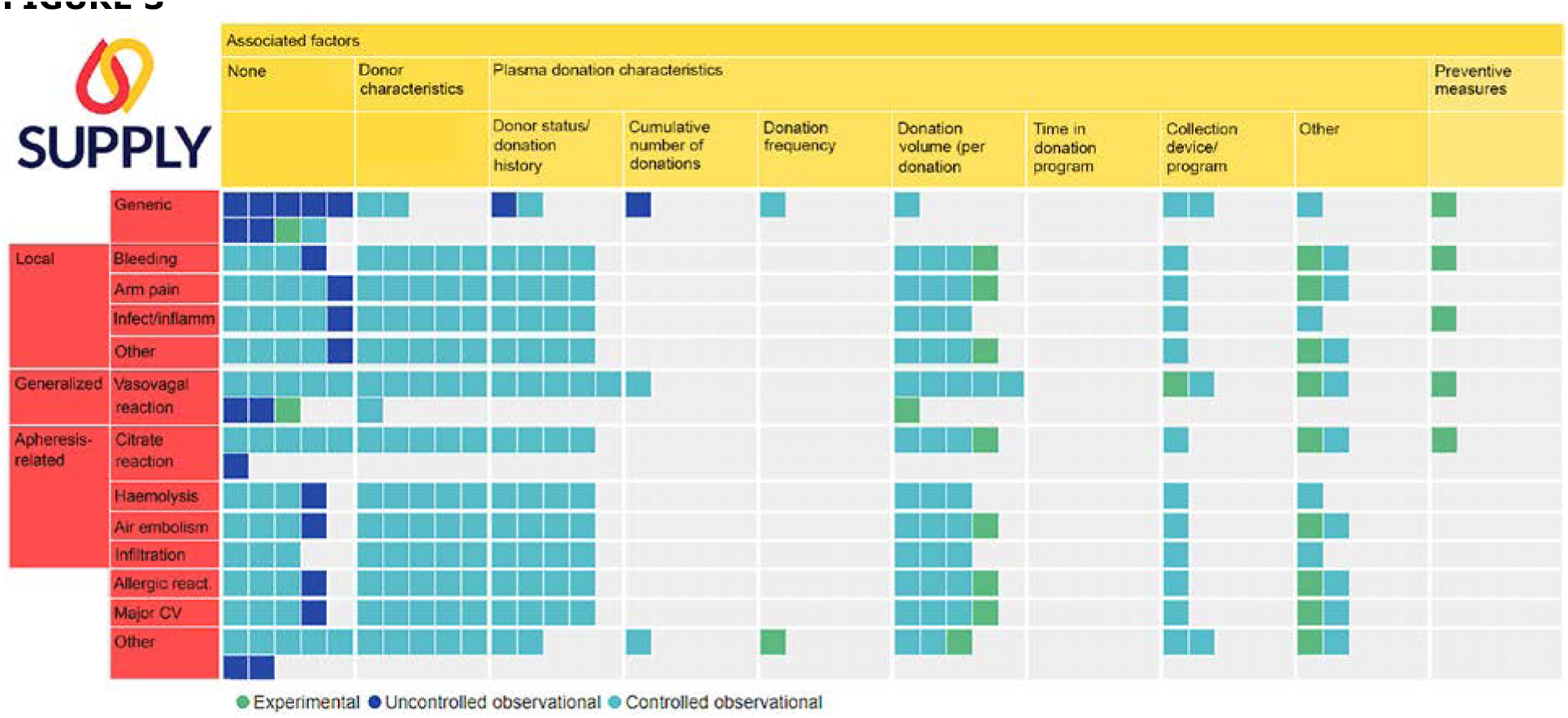
Evidence gap map with adverse events. The columns contain factors that have been studied in association with adverse events. Squares represent studies, subdivided according to study design. An interactive version of the map is available <u>online</u>, in which descriptions of categories can be obtained by hovering over the column/row headers, lists of studies within each category can be obtained by clicking on the map, and studies can be filtered based on study design, publication type, publication date, location, publication language, follow-up period, population, and whether or not convalescent plasma was donated. Generated using v.2.2.4 of EPPI-Mapper powered by EPPI Reviewer and created by the Digital Solution Foundry team.

**FIGURE 4.**
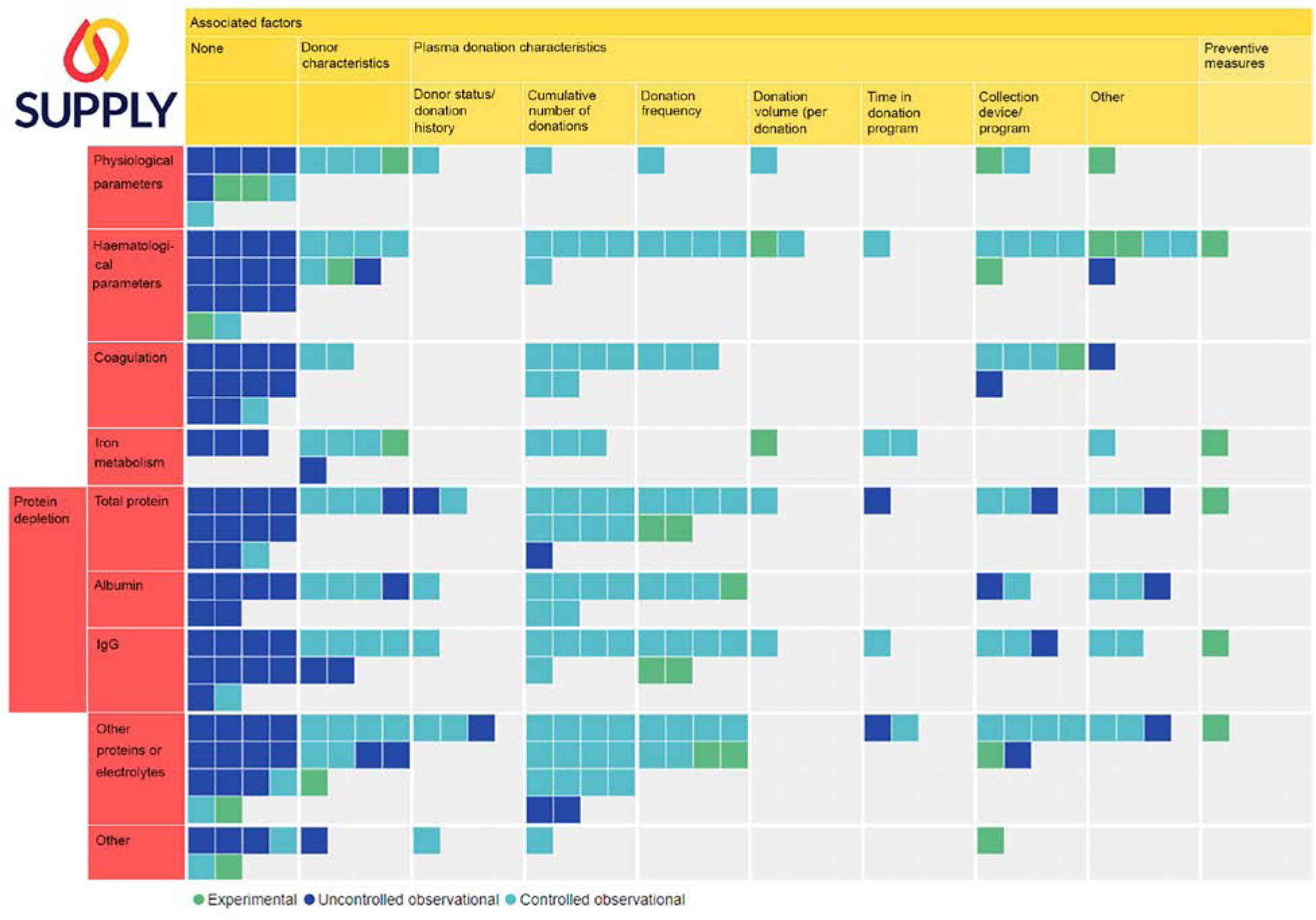
Evidence gap map with health effects. The columns contain factors that have been studied in association with health effects. Squares represent studies, subdivided according to study design. An interactive version of the map is available <u>online</u>, in which descriptions of categories can be obtained by hovering over the column/row headers, lists of studies within each category can be obtained by clicking on the map, and studies can be filtered based on study design, publication type, publication date, location, publication language, follow-up period, population, and whether or not convalescent plasma was donated. Generated using v.2.2.4 of EPPI-Mapper powered by EPPI Reviewer and created by the Digital Solution Foundry team.

#### Population

Most studies (68 out of 99) exclusively included **plasmapheresis donors**, whereas others included an additional, separately analysed, group of blood donors, plateletpheresis or other types of apheresis donors (red or white blood cells) (see <u>EGM</u>) with population filter). Five studies included **convalescent** plasma donors.

#### Publication date, location, and language

Research articles were published between 1956 and October 2022 and registrations between 2016 and 2022. Around 55% of the studies was performed in **Europe** (25 in Germany, 6 in Croatia, 5 in Italy, 3 in France, 3 in Poland, 2 in Denmark, 2 in Norway, 2 in UK, 1 in Russia, Slovakia, Sweden, Switzerland, The Netherlands, and Ukraine each), followed by **North America** (21 in the USA, 6 in Canada, 2 in Cuba), 5 in **Asia** (2 in Japan, 1 in China, India, and Iran each), and 4 in **Australia**.

Two research articles contained data from different countries, including Australia, Brazil, The Netherlands, Wales, the USA, and Singapore or Europe and the USA. Registrations were from Australia (n = 2), Czech Republic, the USA, or Norway. Eighteen studies were not in English.

#### Study design

Most published studies (n = 88) had an **observational** design, of which 56 were **controlled** studies (including 41 cohorts (26 retrospective, 8 prospective, and 7 unclear), 13 controlled before-and-after, 1 case-control, and 1 cross-sectional study) and 32 were **uncontrolled**, monitoring a group of donors before and after donation(s) (n =19) or only after donation(s) (n = 13). Thirteen studies had a **controlled experimental** design (i.e., (non-)randomized-controlled trials), comparing plasmapheresis donors to a no-donation control group ^20, 21^, or blood donors ^18, 22^; or investigating the effect of iron supplementation ^23^, saline infusion ^24–26^, apheresis device or program ^19, 27, 28^, donation volume ^29^, or donation frequency ^30^. Of note, research articles using two different study designs depending on the factor or outcome under study, were counted twice here and in the EGM ^26, 31–36^.

Registrations were classified as controlled experimental (n = 3), controlled observational (n = 1), or uncontrolled observational (n = 1).

### Reported outcomes

Thirty-five research articles and 3 registrations mentioned AE assessment, while 74 research articles and 3 registrations mentioned the investigation of other health effects in plasmapheresis donors (Fig. 2).

#### Adverse events

Of the 35 articles, over 40% mentioned the measurement of AEs without specifying the type of events, adopted surveillance tool, or categorization (classified as ’generic’ in the <u>EGM</u>) (Fig. 3). Assessed AEs included vasovagal reactions (n = 17), complications with local symptoms (directly caused by needle insertion) (n = 14), apheresis-related events (n = 14), allergic reactions (n = 11), major cardiovascular events (n = 11), or other (n = 14; including fractures, non-localized infections, or AEs classified as ’other’ according to the adopted vigilance scheme). The ’Standard for Surveillance of Complications Related to Blood Donation’ ^8^ was the most commonly used tool ^37-^ 43.

About half of the articles (n = 18) simply assessed AE frequency ^18, 32–34, 39, 41, 43–54^, whereas the other half (n = 17) investigated plasmapheresis-related factors that may be associated to the occurrence of AEs (in order of frequency):

- donor characteristics (age, sex, body weight, body mass index, or pre-donation values of blood pressure, pulse, blood volume, or IgG, total protein, haemoglobin (Hb), haematocrit (Hct) levels) 35,37,38,42,55-59,

- donor status (first time versus repeat) ^37, 38, 42, 55, 57, 59, 60^,

- donation volume per session ^24, 31, 37, 38, 57, 59, 61^,

- plasma collection device or program ^19, 35, 36, 58^,

- cumulative number of donations ^56, 57, 60^,

- donation frequency ^61^,

- or other factors (extracorporeal blood volume ^62^, geographical region of plasma collection^40^, or donation with or without saline infusion ^24^).

Three registrations aim to assess the effect of donation frequency ^63^, applied muscle tension ^64^ (registration withdrawn), or did not include any plasmapheresis-related factors ^65^.

#### Health effects

Out of 73 studies assessing health effects, the majority investigated protein depletion (total protein, albumin, and/or IgG levels) (n = 40) and/or haematological parameters (full blood count, Hct, Hb, lymphocyte, platelet, red blood cell, reticulocyte, schizocyte, and/or white blood cell counts) (n = 37) (Fig. 4). Others included coagulation-related outcomes (n = 24), physiological parameters (n = 13), iron metabolism (n = 12), other proteins/electrolytes measured in blood/plasma (n = 45; a list of which is available on our OSF page ^7^ at https://osf.io/5nz3v), and/or products used during plasmapheresis (citrate, MEHP, DEHP) or synthetics (n = 9).

Twenty-eight studies simply measured health effects ^18, 20, 22, 31, 35, 36, 45–47, 50–52, 66–81^, whereas most studies investigated plasmapheresis-related factors that may be associated with the occurrence of health effects, including (in order of frequency):

- donor characteristics (age, sex, body weight, body mass index, or pre-donation values of blood pressure, pulse, blood volume, or IgG, total protein, Hb, Hct levels) ^21, 29, 33, 58, 82–95^,

- cumulative number of donations ^33, 88, 89, 92, 93, 95–105^,

- donation frequency ^30,33,61,90,95,98-102^,

- plasma collection device or program ^27, 28, 32, 36, 58, 106–109^,

- donor status (first time versus repeat) ^68, 88, 90, 110, 111^,

- donation volume per session ^29, 31, 61, 65^,

- time in donation program ^34, 88, 92^,

- iron supplementation as a preventive measure ^23^,

- or other factors (saline infusion ^25, 26^, compensated, paid, or unpaid donors ^112^, source plasma versus RhIg plasma ^113^, the anticoagulant used ^114^).

Three identified registrations mentioned the following factors: collection device and left versus right antecubital donation ^115^, cumulative number of donations (i.e., 4 vs 8 cycles) ^116^, or donation frequency ^63^.

### Factors associated with adverse events and/or health effects

The effectiveness of preventive measures, donor status (first-time versus repeat), cumulative number of donations, and donation frequency in relation to donor health, were a priori identified as SUPPLY project objectives ^4^, and are elaborated upon below.

#### The effects of preventive measures on donor safety/health

A randomized controlled trial assessed the effect of **daily iron supplementation** (versus placebo) on haematological parameters, iron metabolism, protein depletion, alanine aminotransferase, and viral markers in menstruating women who donated plasma at one-week intervals over 24 weeks ^23^. An RCT registration on the effect of **applied muscle tension** on AE rates has been withdrawn ^64^.

#### Donor safety/health in first-time versus repeat plasmapheresis donors

Ten observational studies assessed AEs ^37, 38, 42, 55, 57, 59, 60^ or health effects ^68, 88, 111^ in first-time and repeat plasmapheresis donors separately. Half of these studies were published recently (after 2020) ^37, 38, 42, 57, 59^. Assessed AEs included ’all AEs’ (generic) ^55, 60^, vasovagal reactions ^37, 38, 42, 57, 59^, local symptoms, apheresis-related complications, allergic reactions, major cardiovascular events, and other types ^37, 38, 42, 59^. Health-related outcomes included total protein ^88, 111^, albumin ^111^, IgG and other Ig levels ^111^, monoclonal gammopathies or other gamma globulin abnormalities ^88^, or DEHP (plasticizer) in donor plasma or blood ^68^.

#### The association between cumulative number of donations and donor safety and/or health

Twenty observational studies investigated the association between the cumulative number of plasmaphereses (i.e., multiple donations) and donor safety and/or health. No experimental studies were identified.

Three studies reported on AEs ^57, 60, 117^, including ’all AEs’ (see ’generic’ in EGM) ^60^, vasovagal reactions ^57^, or (osteoporotic) fractures ^56^. In addition, 18 studies assessed health markers measured in donor blood or plasma, including – in order of frequency – total protein levels ^88, 89, 95-^ 97,101,102,104,105, albumin 89,95,97,101,102,104, IgG levels 95,96,101,102,104, coagulation tests 99,102-105, haematological parameters ^95, 98, 99, 104, 105^, iron metabolism ^92, 93, 104^, or other ^33, 88, 89, 95–98, 100–105^. One study addressed physiological parameters ^90^.

Finally, a case series registration aims to investigate indicators of biological age after 4 versus 8 plasmaphereses ^116^.

#### The association between donation frequency and donor safety and/or health

Studies are described below if (1) they specified the time interval between subsequent donations or the number of donations per given time unit (e.g., weekly donation, bi-weekly donation, 3 donations every 2 weeks) and (2) if the donation frequency was constant/unchanged during the study period. For example, studies providing the number of donations per year are not included under this category if it was unclear whether the donation frequency remained constant throughout the year.

Ten studies, including 1 experimental study and 9 controlled observational studies, investigated the association between frequency of plasmaphereses and donor safety and/or health. The most recent article dates from 2015, and 70% were published before 2000. Five studies took place within the same research group in Croatia ^98–102^.

One study assessed adverse events (see ’generic’ in EGM) ^61^, whereas all 10 studies assessed the impact on donor health, including – in order of frequency – total protein ^30, 61, 95, 101, 102^, IgG levels ^30, 61, 95, 101, 102^, albumin ^30, 95, 101, 102^, haematological parameters ^61, 95, 98, 99^, coagulation tests ^98, 99, 102^, or other health markers measured in donor blood or plasma ^30, 33, 95, 98, 99, 101, 102^. One study included physiological parameters ^90^.

Finally, an RCT registration aims to investigate the effect of donation frequency on total protein, IgG levels, other plasma proteins, and psychological distress ^63^.

#### Long-term (≥1 week) follow-up of plasmapheresis donors

Thirty-eight studies (incl. 3 registrations) had a follow-up period (i.e., time between the first donation and last donor assessment) of 1 week or longer. Studies are not included in the overview below if they assessed the effect of multiple donations without specifying (or allowing calculation of) the follow-up period ^26^.

#### Follow-up after a single donation (n = 6)

Five studies (and 1 registration ^65^) investigated the effect of a single donation on AEs or health effects and included a follow-up period of 1 week ^22, 65, 107^, 3 weeks ^87^, 1 month after the donation^38^, or unspecified (presumably longer than 1 week) ^41^.

#### Follow-up during or after multiple donations (n = 32)

Thirty-two studies monitored donors (before and) after or during a specified period in which multiple donations were given. Studies are subdivided according to study design and listed in order of increasing follow-up period.

#### Controlled experimental studies (n = 6)

Three controlled experimental studies (and 1 registration ^63^) examined the effect of iron supplementation ^23^, donation frequency ^30, 63^, or collection program ^19^ on donor health with a follow-up period of **1-6 months**. Finally, 2 studies had a longer follow-up period (**6-12 months**), comparing whole blood donations, plasma donations, and observation only ^18^; or different donation volumes ^29^.

#### Controlled observational studies (n = 16)

Three studies assessed donors who frequently donated for **1-6 months**, monitoring AEs and haematological parameters after 10 weekly donations ^36^, serum protein levels before and after 5 monthly donations ^96^, or blood pressure during 4 months (prospective study) ^90^. Five retrospective studies investigated donors who frequently underwent plasmapheresis for **1 year**, focusing on either iron metabolism ^86, 92, 93, 104^, haematological parameters ^91, 104^, or other proteins ^104^. Eight papers reported a study period of **more than 1 year** and up to 23 years, looking at a wide variety of outcomes and including 4 prospective studies ^35, 61, 71, 94^, 3 retrospective cohorts ^52, 56, 102^, and 1 cohort with unclear classification ^34^.

#### Uncontrolled observational studies (n = 10)

Three uncontrolled before-and-after studies assessed health effects with a study period of **1-6 months** ^36, 66, 74^. Two case series, one of which is a registration ^116^, mentioned a **6-to-12-month** follow-up for health effects ^79^. Finally, we identified 5 studies with a follow-up period of **over 1 year**: 1 before-and-after study assessing AEs ^47^ and 4 case series looking at other health effects^35, 73, 78, 88^.

## Discussion

The current scoping review identified 94 research articles and 5 registrations focusing on AEs and/or other health effects in plasmapheresis donors, most of which were performed in **Europe** (55%) or the **USA** (20%). The majority of studies (n = 91, incl. 2 registrations) employed an **observational design** (57 controlled and 33 uncontrolled, monitoring a group of donors (before and) after donation(s)). Only 16 studies (incl. 3 registrations) had an **experimental design**. Given that we did not identify any systematic review (registration), our ongoing systematic review on donation frequency ^11^ that was performed in follow-up of the current scoping review, is presumably the first one to focus on the impact of plasmapheresis on donor health.

In the framework of developing safety guidelines for plasmapheresis donors while increasing plasma collection within Europe, it is of paramount importance to consider the **effect of frequent and long-term plasmapheresis donation**. Although we identified 10 studies investigating the association between donation frequency and donor health, there was only one experimental study^30^. In addition, studies are relatively old (70% were published before 2000 and the most recent one dates from 2015), and 5 studies are from the same research group. More than 30 studies analysed donors during or after a specified period (1 month to 23 years) in which multiple donations were given. However, 10 of these studies did not include a control group, limiting the certainty of their conclusions. Among the controlled studies, the majority were observational (n = 16) and retrospective (n = 11). Such retrospective studies are subject to bias, given that long-term donors are self-selected to withstand the donation frequency under evaluation.

Studies on **preventive interventions** against AEs or other health effects are scarce, given that there is only one such study, reporting on the effect of iron supplementation. The effectiveness of identified interventions in blood donors (e.g., to reduce vasovagal reactions ^118^) need to be validated in plasmapheresis donors.

A strength of the current scoping review includes its **rigorous and systematic methodology**, as reflected by the independent screening and extraction conducted by 2 reviewers. This review covers a **broad research area**, encompassing all studies that recorded AEs and other health-related outcomes in plasmapheresis donors. In addition to a detailed table (https://osf.io/h8js5) with study characteristics, a comprehensive, accessible, and interactive visual overview is provided in the form of an <u>EGM</u>.

On the other hand, a first limitation of this scoping review is the **arbitrary and ambiguous nature of the developed factor/outcome categories**, even though these categories were developed in consultation with content experts (SM, KvdH, CE, PT, VC). To ensure transparency, descriptions, and examples of these categories can be obtained by hovering over their labels in the data charting table (https://osf.io/h8js5) and <u>EGM</u>. For further clarification, lists of all encountered factors (https://osf.io/uv6fe) and health effects (https://osf.io/5nz3v) are available on our OSF page, with each of the factors/outcomes listed under their relevant category. Second, it should be noted that scoping reviews typically do not involve extracting study results and judging the quality of studies, and therefore **do not contribute to the development of practical recommendations** regarding donor safety. Rather, the scoping review provides an overview of the available evidence and evidence gaps for researchers, blood banks, plasma industry representatives, and policy-makers.

To conclude, based on the evidence (gaps) identified in the current scoping review, we propose the following research recommendations. First, additional experimental studies are required to investigate the **impact of various donation frequencies on donor health**, such as the registered study by Strand et al. ^63^. The results of these studies will help establish a safe upper limit for plasma donation frequency. Second, to obtain conclusive evidence regarding the health effects of frequent and long-term donation, more **controlled prospective studies** are warranted. Ideally, these studies would monitor drop-out rates and reasons, as well as the long-term health of those who have dropped out; and incorporate intention-to-treat analyses to mitigate potential bias arising from the healthy donor effect ^119^. Finally, considering the incomplete **reporting on AEs** in several identified studies, we recommend that future studies specify the exact timing of outcome measurement, and describe the adopted surveillance tools or AEs that are being monitored.

## Funding

This work was funded by the EU4Health Programme by the European Union (101056988) and the Foundation for Scientific Research of the Belgian Red Cross.

## Conflict of interest

HVR, NS, VC, EBD, and TD are employed by Belgian Red Cross–Flanders, responsible and reimbursed for supplying adequate quantities of safe blood products to hospitals in Flanders and Brussels. PT is employed by the Etablissement Français du Sang, the French transfusion public service in charge of blood, plasma, and platelet collection in France. KvdH is employed by Sanquin, responsible for safe blood supply in the Netherlands. The authors have disclosed no conflicts of interest.

## Contributions

NS: Reviewer 1, Writing – original draft; HVR: Reviewer 2, Project coordination, Writing – review & editing, Supervision; TD: Reviewer 3, Writing – review & editing; SM: Writing – review & editing, Validation; KVDH/VC/CE/PT: Conceptualization, Writing – review & editing, Validation, Funding acquisition; EDB: Supervision, Writing – review & editing.

## Data Availability

All data produced are available online at https://osf.io/kbv6z.

## Notes

### Competing Interest Statement

The authors have declared no competing interest.

### Summary of Updates

No content changes have been made to the main text. 1. Embedded links were not working in the previous version, which has been fixed, 2. COI and contributions for EBD were added, 3. Font was altered to enhance readability.

