## Appendix 2 for "Safety and protection of plasma donors: A scoping review and evidence (gap) map"

### Appendix 2: Search Strategy

**MEDLINE and PubMed Central (PubMed interface)**

**1. Population** (plasma donor)
"plasma donor*"[TIAB] OR "plasma donat*"[TIAB]
OR (("Plasmapheresis"[Mesh:NoExp] OR "plasmapheres*"[TIAB] OR "plasmaphaeres*"[TIAB] OR "plasma collection"[TIAB] OR "plasma withdrawal"[TIAB] OR "plasma removal"[TIAB]
OR "Blood component removal"[Mesh:NoExp] OR "apheres*"[TIAB] OR "aphaeres*"[TIAB] OR "pheres*"[TIAB] OR "phaeres*"[TIAB]
OR "Plateletpheresis"[Mesh:NoExp] OR "plateletpheres*"[TIAB] OR "plateletphaeres*"[TIAB] OR thrombocytapheres*[TIAB] OR thrombocytopheres*[TIAB] OR thrombapheres*[TIAB] OR thromboapheres*[TIAB] OR thrombophores*[TIAB] OR thrombocytaphaeres*[TIAB] OR thrombocytophaeres*[TIAB] OR thrombaphaeres*[TIAB] OR thromboaphaeres*[TIAB])
AND ("donor*"[TIAB] OR "donat*"[TIAB]))

**2. Concept** (safety)
"safety"[MESH] OR "safe*"[TIAB] OR "efficacy"[TIAB] OR "effect*"[TIAB]
OR (("adverse"[TIAB] OR "undesirable"[TIAB] OR "side"[TIAB] OR "acute"[TIAB] OR "short-term"[TIAB] OR "long-term"[TIAB] OR "shortterm"[TIAB] OR "longterm"[TIAB] OR vital[TIAB]) AND ("reaction*"[TIAB] OR "consequence*"[TIAB] OR "event*"[TIAB] OR "outcome*"[TIAB] OR "symptom*"[TIAB]))
OR "tolerability"[TIAB] OR "harm"[TIAB] OR "complication*"[TIAB]
OR "plasmavigilance"[TIAB] OR "hemovigilance"[TIAB] OR "haemovigilance"[TIAB]
OR "donor vigilance"[TIAB]
OR "health"[TIAB] OR "monitor*"[TIAB] OR "surveillance"[TIAB]
OR "Plasmapheresis/adverse effects"[MESH] OR "Blood Component Removal/adverse effects"[Mesh] OR "Plateletpheresis/adverse effects"[Mesh] OR "Monitoring, physiologic"[Mesh]

**3. Concept** (risk and prevention)
"Preventive Medicine"[Mesh] OR "prevention and control"[Subheading] OR "Health Promotion"[Mesh] OR "Risk Factors"[Mesh] OR "prevent*"[TIAB] OR "protect*"[TIAB] OR "risk factor"[TIAB] OR "risk factors"[TIAB] OR "Risk Assessment"[Mesh:NoExp] OR "Risk"[Mesh:NoExp] OR "donor characteristic*"[TIAB]
OR "Plasmapheresis/methods"[MESH]

#1 AND (#2 OR #3)

**Embase (Embase.com)**

**1. Population** (plasma donor)
'plasma donor*':ti,ab OR 'plasma donat*':ti,ab
OR ((Plasmapheresis/de OR plasmapheres*:ti,ab OR plasmaphaeres*:ti,ab OR 'plasma collection':ti,ab OR 'plasma withdrawal':ti,ab OR 'plasma removal':ti,ab
OR apheresis/de OR apheres*:ti,ab OR aphaeres*:ti,ab OR pheres*:ti,ab OR phaeres*:ti,ab
OR thrombocytopheresis/de OR plateletpheres*:ti,ab OR plateletphaeres*:ti,ab OR thrombocytapheres*:ti,ab OR thrombocytopheres*:ti,ab OR thrombapheres*:ti,ab OR thromboapheres*:ti,ab OR thrombophores*:ti,ab OR thrombocytaphaeres*:ti,ab OR thrombocytophaeres*:ti,ab OR thrombaphaeres*:ti,ab OR thromboaphaeres*:ti,ab) AND (donor*:ti,ab OR donat*:ti,ab))

**2. Concept** (safety)
safety/exp OR safe*:ti,ab OR efficacy:ti,ab OR effect*:ti,ab
OR ((adverse:ti,ab OR undesirable:ti,ab OR side:ti,ab OR acute:ti,ab OR 'short-term':ti,ab OR 'long-term':ti,ab OR shortterm:ti,ab OR longterm:ti,ab OR vital:ti,ab) AND (reaction*:ti,ab OR consequence:ti,ab OR event*:ti,ab OR outcome*:ti,ab OR symptom*:ti,ab))
OR tolerability:ti,ab OR harm:ti,ab OR complication*:ti,ab
OR plasmavigilance:ti,ab OR hemovigilance:ti,ab OR haemovigilance:ti,ab
OR 'donor vigilance':ti,ab
OR health:ti,ab OR monitor*:ti,ab OR surveillance:ti,ab
OR 'physiologic monitoring'/exp

**3. Concept** (risk and prevention)

'Preventive Medicine'/exp OR 'Health Promotion'/exp OR 'Risk Factor'/exp OR prevent*:ti,ab OR protect*:ti,ab OR 'risk factor':ti,ab OR 'risk factors':ti,ab OR 'Risk Assessment'/de OR 'Risk'/de OR 'donor characteristic*':ti,ab

#1 AND (#2 OR #3)

**Cochrane Library (systematic reviews and controlled trials)**

**1. Population** (plasma donor)
(plasma NEXT donor*):ti,ab,kw OR (plasma NEXT donat*):ti,ab,kw
OR (([mh ^"Plasmapheresis"] OR plasmapheres*:ti,ab,kw OR plasmaphaeres*:ti,ab,kw OR "plasma collection":ti,ab,kw OR "plasma withdrawal":ti,ab,kw OR "plasma removal":ti,ab,kw
OR [mh ^"Blood component removal"] OR apheres*:ti,ab,kw OR aphaeres*:ti,ab,kw OR pheres*:ti,ab,kw OR phaeres*:ti,ab,kw
OR [mh ^"Plateletpheresis"] OR plateletpheres*:ti,ab,kw OR plateletphaeres*:ti,ab,kw OR thrombocytapheres*:ti,ab,kw OR thrombocytopheres*:ti,ab,kw OR thrombapheres*:ti,ab,kw OR thromboapheres*:ti,ab,kw OR thrombophores*:ti,ab,kw OR thrombocytaphaeres*:ti,ab,kw OR thrombocytophaeres*:ti,ab,kw OR thrombaphaeres*:ti,ab,kw OR thromboaphaeres*:ti,ab,kw)
AND (donor*:ti,ab,kw OR donat*:ti,ab,kw))

**2. Concept** (safety)
[mh "safety"] OR safe*:ti,ab,kw OR efficacy:ti,ab,kw OR effect*:ti,ab,kw
OR ((adverse:ti,ab,kw OR undesirable:ti,ab,kw OR side:ti,ab,kw OR acute:ti,ab,kw OR "short-term":ti,ab,kw OR "long-term":ti,ab,kw OR shortterm:ti,ab,kw OR longterm:ti,ab,kw OR vital:ti,ab,kw) AND (reaction*:ti,ab,kw OR consequence:ti,ab,kw
OR event*:ti,ab,kw OR outcome*:ti,ab,kw OR symptom*:ti,ab,kw))
OR tolerability:ti,ab,kw OR harm:ti,ab,kw OR complication*:ti,ab,kw
OR plasmavigilance:ti,ab,kw OR hemovigilance:ti,ab,kw OR haemovigilance:ti,ab,kw
OR "donor vigilance":ti,ab,kw
OR health:ti,ab,kw OR monitor*:ti,ab,kw OR surveillance:ti,ab,kw
OR [mh "Monitoring, Physiologic "]

**3. Concept** (risk and prevention)
[mh "Preventive Medicine"] OR [mh "Health Promotion"] OR [mh "Risk Factors"] OR prevent*:ti,ab,kw OR protect*:ti,ab,kw OR (risk NEXT (factor OR factors)):ti,ab,kw OR [mh ^"Risk Assessment"] OR [mh ^"Risk"] OR (donor NEXT characteristic*):ti,ab,kw

#1 AND (#2 OR #3)

**Web of Science Core Collection (Science Citation Index Expanded (SCI-EXPANDED) and Conference Proceedings Citation Index- Science (CPCI-S))**

**1. Population** (plasma donor)
TS=("plasma donor*") OR TS=("plasma donat*")
OR ((TS=("plasmapheres*") OR TS=("plasmaphaeres*") OR TS=("plasma collection") OR TS=("plasma withdrawal") OR TS=("plasma removal")
OR (TS="apheres*") OR (TS="aphaeres*") OR (TS="pheres*") OR (TS="phaeres*")
OR (TS="plateletpheres*") OR (TS="plateletphaeres*") OR (TS="thrombocytapheres*") OR (TS="thrombocytopheres*") OR (TS="thrombocytepheres*") OR (TS="thrombapheres*") OR (TS="thromboapheres *") OR (TS="thrombophores*") OR (TS="thrombocytaphaeres*") OR (TS="thrombocytophaeres*")
OR (TS="thrombocytephaeres*") OR (TS="thrombaphaeres*") OR (TS="thromboaphaeres*"))
AND (TS=("donor*") OR TS=("donat*")))

**2. Concept** (safety)
TS=("safe*") OR TS=("efficacy") OR TS=("effect*")
OR ((TS=("adverse") OR TS=("undesirable") OR TS=("side") OR TS=("acute") OR TS=("short-term") OR TS=("long-term") OR TS=("shortterm") OR TS=("longterm") OR TS=("vital")) AND (TS=("reaction*") OR TS=("consequence*") OR TS=("event*") OR TS=("outcome*") OR TS=("symptom*")))
OR TS=("tolerability") OR TS=("harm") OR TS=("complication*")
OR TS=("plasmavigilance") OR TS=("hemovigilance") OR TS=("haemovigilance")
OR TS=("donor vigilance")
OR TS=("health") OR TS=("monitor*") OR TS=("surveillance")

**3. Concept** (risk and prevention)
TS=("prevent*") OR TS=("protect*") OR TS=("risk factor") OR TS=("risk factors") OR TS=("donor characteristic*")

#1 AND (#2 OR #3)

**CINAHL (Ebsco interface)**

**1. Population** (plasma donor)
(TI "plasma donor*" OR AB "plasma donor*") OR (TI "plasma donat*" OR AB "plasma donat*")
OR (((MH "Plasmapheresis") OR (TI "plasmapheres*" OR AB "plasmapheres*") OR (TI "plasma-phaeres*" OR AB "plasmaphaeres*") OR (TI "plasma collection" OR AB "plasma collection") OR (TI "plasma withdrawal" OR AB "plasma withdrawal") OR (TI "plasma removal" OR AB "plasma removal")
OR (MH "Blood component removal") OR (TI "apheres*" OR AB "apheres*") OR (TI "aphaeres*" OR AB "aphaeres*") OR (TI "pheres*" OR AB "pheres*") OR (TI "phaeres*" OR AB "phaeres*")
OR (MH "Plateletpheresis") OR (TI "plateletpheres*" OR AB "plateletpheres*") OR (TI "plateletphaeres*" OR AB "plateletphaeres*") OR (TI "thrombocytapheres*" OR AB "thrombocytapheres*") OR (TI "thrombocytopheres*" OR AB "thrombocytopheres*") OR (TI "thrombapheres*" OR AB "thrombapheres*") OR (TI "thromboapheres*" OR AB "thromboapheres*") OR (TI "thrombophores*" OR AB "thrombophores*") OR (TI "thrombocytaphaeres*" OR AB "thrombocytaphaeres*") OR (TI "thrombocytophaeres*" OR AB "thrombocytophaeres*") OR (TI "thrombaphaeres*" OR AB "thrombaphaeres*") OR (TI "thromboaphaeres*" OR AB "thromboaphaeres*"))
AND ((TI "donor*" OR AB "donor*") OR (TI "donat*" OR AB "donat*")))

**2. Concept** (safety)
(MH safety+) OR (TI safe* OR AB safe*) OR (TI effect* OR AB effect*)
OR (((TI adverse OR AB adverse) OR (TI undesirable OR AB undesirable) OR (TI side OR AB side) OR (TI acute OR AB acute) OR (TI short-term OR AB short-term) OR (TI long-term OR AB long-term) OR (TI shortterm OR AB shortterm) OR (TI long-term OR AB longterm)
OR (TI vital OR AB vital)) AND ((TI reaction* OR AB reaction*) OR (TI consequence OR AB consequence) OR (TI event* OR AB event*) OR (TI outcome* OR AB outcome*) OR (TI symptom* OR AB symptom*)))
OR (TI tolerability OR AB tolerability) OR (TI harm OR AB harm) OR (TI complication* OR AB complication*)
OR (TI plasmavigilance OR AB plasmavigilance) OR (TI hemovigilance OR AB hemovigilance) OR (TI haemovigilance OR AB haemovigilance)
OR (TI "donor vigilance" OR AB "donor vigilance")
OR (TI health OR AB health) OR (TI monitor* OR AB monitor*) OR (TI surveillance OR AB surveillance)
OR (MH "Monitoring, physiologic"+)

**3. Concept** (risk and prevention)

(MH "Preventive Medicine"+)
OR "prevention and control[Subheading]" OR (MH "Risk Factors"+) OR (TI prevent* OR AB prevent*) OR (TI protect* OR AB protect*) OR (TI "risk factor" OR AB "risk factor") OR (TI "risk factors" OR AB "risk factors")OR (MH "Risk Assessment")  OR (TI "donor characteristic*" OR AB "donor characteristic*")

#1 AND (#2 OR #3)

**Transfusion Evidence Library**
Filter: Clinical speciality > Blood Donors
((plasma OR plasmapheresis OR plasmaphaeresis OR apheresis OR aphaeresis OR pheresis OR phaeresis OR plateletpheresis OR plateletphaeresis OR thrombocytapheresis OR thrombocytopheresis OR thrombapheresis OR thromboapheresis OR thrombophoresis OR thrombocytaphaeresis OR thrombocytophaeresis OR thrombaphaeresis OR thromboaphaeresis) AND (donor OR donors OR donation OR donations))

**PROSPERO International Prospective Register of Systematic Reviews**
((plasma OR plasmapheresis OR plasmaphaeresisOR apheresis OR aphaeresis OR pheresis OR phaeresis OR plateletpheresis OR plateletphaeresis OR thrombocytapheresis OR thrombocytopheresis OR thrombapheresis OR thromboapheresis OR thrombophoresis OR thrombocytaphaeresis OR thrombocytophaeresis OR thrombaphaeresis OR thromboaphaeresis) AND (donor OR donors OR donation OR donations))
AND (safe OR safety OR effect OR ((adverse OR undesirable OR side OR acute OR "short-term" OR "long-term" OR shortterm OR longterm) AND (reaction OR event OR outcome OR symptom OR reactions OR effects OR events OR outcomes OR symptoms)) OR tolerability OR harm OR complication OR complications OR plasmavigilance OR hemovigilance OR haemovigilance OR "donor vigilance" OR health OR monitor OR monitoring OR surveillance OR prevent OR prevention OR protect OR protection OR risk OR characteristic OR characteristics)

**International Clinical Trials Registry Platform (trialsearch.who.int/)**
((plasma OR plasmapheresis OR plasmaphaeresis OR apheresis OR aphaeresis OR pheresis OR phaeresis OR plateletpheresis OR plateletphaeresis OR thrombocytapheresis OR thrombocytopheresis OR thrombapheresis OR thromboapheresis OR thrombophoresis OR thrombocytaphaeresis OR thrombocytophaeresis OR thrombaphaeresis OR thromboaphaeresis) AND (donor OR donors OR donation OR donations))
AND (safe OR safety OR effect OR ((adverse OR undesirable OR side OR acute OR "short-term" OR "long-term" OR shortterm OR longterm) AND (reaction OR event OR outcome OR symptom OR reactions OR effects OR events OR outcomes OR symptoms)) OR tolerability OR harm OR complication OR complications OR plasmavigilance OR hemovigilance OR haemovigilance OR "donor vigilance" OR health OR monitor OR monitoring OR surveillance OR prevent OR prevention OR protect OR protection OR risk OR characteristic OR characteristics)

**Clinicaltrials.gov** (split up into 3 searches due to character limit) search in 'Other terms' field
1)
(((plasma OR plasmapheresis OR apheresis OR plateletpheresis) AND (donor OR donation)))
AND
(safe OR safety OR effect OR ((adverse OR undesirable OR side OR acute OR "short-term" OR "long-term") AND (reaction OR event OR outcome OR symptom)))

2)
(((plasma OR plasmapheresis OR apheresis OR plateletpheresis) AND (donor OR donation)))
AND

(plasmavigilance OR hemovigilance OR haemovigilance OR "donor vigilance" OR monitor OR monitoring OR surveillance)

3)
(((plasma OR plasmapheresis OR apheresis OR plateletpheresis) AND (donor OR donation)))
AND
(tolerability OR harm OR complication OR health OR prevent OR prevention OR protect OR protection OR risk OR characteristic)
